# Dynamic Modeling of Reported COVID-19 Cases and Deaths with Continuously Varying Case Fatality and Transmission Rate Functions

**DOI:** 10.1101/2020.09.25.20201905

**Authors:** Mingdong Lyu, Randolph Hall

## Abstract

In this paper, we propose an enhanced SEIRD (Susceptible-Exposed-Infectious-Recovered-Death) model with time varying case fatality and transmission rates for confirmed cases and deaths from COVID-19. We show that when case fatalities and transmission rates are represented by simple Sigmoid functions, historical cases and fatalities can be fit with a relative-root-mean-squared-error accuracy on the order of 2% for most American states over the period from initial cases to July 28 (2020). We find that the model is most accurate for states that so far had not shown signs of multiple waves of the disease (such as New York), and least accurate for states where transmission rates increased after initially declining (such as Hawaii). For such states, we propose an alternate multi-phase model. Both the base model and multi-phase model provide a way to explain historical reported cases and deaths with a small set of parameters, which in the future can enable analyses of uncertainty and variations in disease progression across regions.

## Introduction

COVID-19 has challenged the world to react to a new contagious virus in the absence of effective medical treatment and vaccines. Over the course of nine months from the outbreak in December 2019, when the first cases were confirmed in Wuhan, China, until April 24th 2020, 213 countries and territories reported nearly 28 million confirmed cases and a death toll exceeding 900,000 persons [1]. In the meantime, waiting for effective clinical care and vaccination, countries have reacted to the pandemic by controlling travel, implementing large-scale quarantine and restricting gatherings and contact among people, as well as requiring hygiene measures and screening for possible cases.

Based on recent findings, compared to the global SARS epidemic in 2002 and MERS in 2012, COVID-19 has a relatively long incubation period, with a mean time of 5 days [2]. COVID-19 has also been found to be transmissible while individuals are asymptomatic. Meanwhile, disease severity is widely variable, depending on age, comorbidities, baseline health and access to care. Even those with mild or no symptoms, who are often young adults, may still transmit the disease to others. These factors, combined with limited testing and inconsistency in adherence to public health measures, have made the virus hard to contain. Policy makers are also facing a dilemma, balancing the goal of maintaining economic activity against saving lives through strict measures, while at the same time learning about the effectiveness of interventions and the nature of the disease.

In this paper, we seek to improve understanding of the dynamics of how COVID-19 is spread, utilizing a variation of the Susceptible-Exposed-Infectious-Recovered-Death (SEIRD) model. Our key innovation is representation of the transmission rate and case fatality rate as continuously varying functions, which are optimally fit to historical data on confirmed cases and deaths. We surmise that neither parameter is static, as they are influenced by the enactment and adherence to public health measures and medical care, neither of which is constant over time. We have applied our model to all 50 American states to derive insights into how the disease has spread in different localities, which are influenced by population health, disease exposure, localized public health interventions and messaging, in addition to other place specific factors.

## Prior Research

Prior research on COVID-19 has estimated disease-specific parameters, such as the basic reproduction number and latent period [3–5], demonstrating why the disease is highly transmissible. Mathematical models have also been used to analyze different transmission scenarios, to inform policy makers as to possible futures and the effects of interventions. For example, according to a statistical guideline model published in 2005 [6], the state of New York reacted to the urgent shortage of ventilators by requesting more ventilators from the federal government and implementing new interventions, such as closures of schools and restaurants [7].

Another use of disease transmission models has been to predict and plan for future demands on the health care system, such as demands for hospital beds (ICU in particular) and needs for health care resources, such as ventilators. Toward that goal, [8] provides a statistical model of death data to predict future fatalities, assuming that social distancing measures are maintained. From the projected fatality data, they estimated hospital utilization with an individual-level microsimulation model based on the historical statistics of age-specific ICU admission. [9] simulates the COVID-19 outbreak, parameterized with the US population demographics, with a compartmental model under different scenarios of self-isolation, projecting hospital utilization and recognizing the mitigation effect of self-isolation on hospital capacity.

Due to the limits of testing methods, the long incubation period, and cases with mild or no symptoms and delayed reporting, there is potentially a huge (and unknown) number of unreported cases, the extent to which could affect the future evolution of the epidemic. Some researchers, therefore, have used the SIR (symptomatic-infectious-recovered) model and SEIRD to estimate the number of undetected cases [10] and [11]. Some approaches also incorporate transportation information (such as human migration data and community mobility data) to analyze the impact of travel on disease transmission and thus the effect of travel restriction [12–14]. However, most studies using SEIRD or SIR assume the transmission rate and death rate to be constant over time. With improvement in clinical treatment and changing intervention policies, the transmission rate and fatality rate will be time variable. Therefore, SEIRD models with constant parameters cannot accurately depict the spread of disease. [15] and [16] both consider time dependency of transmission parameters. Godio et al modify the new recovery rate to a sinusoidal function and they adjust the transmission rate according to mobility trends [15]. Piccolomini et al. compare two piecewise time-dependent infection rate functions and fit the infection rate function, incubation rate, and death rate for each uniformly divided time interval [16]. These approaches require introduction of many parameters to depict time dependency, thus risking overfitting and reducing model generality.

In our research we explore use of a concise formulation through which continuously time varying transmission and case fatality rates are modeled with a small number of parameters, which are fit to historical data. Like [7], we utilize a type of logistic function (i.e., a Sigmoid function), but not simply to model reported deaths over time, but to instead model transmission rate and case fatality rate within the SEIRD model. Our objective is to improve the classical SEIRD model through an approach that adapts to the dynamic pattern of transmission under different epidemic scenarios. Thus, we provide insights into transmissibility of the disease while modeling historical data on confirmed cases and confirmed deaths.

## The Proposed Time Varying Model

We draw from the SEIRD compartmental model, which divides the population into five groups: susceptible(*S*), exposed(*E*), infected(*I*), recovered(*R*) and dead(*D*). SEIRD utilizes differential equations to model the evolution of the number of people in these states over time. Susceptible individuals can catch the virus through contact with infectious people and turn into the state of exposed. Exposed people are in a latent state and then progress to the infectious state with a rate inversely related to the incubation period (thus, exposed is defined as a state in which people are not yet infectious). Infected people eventually progress into either the dead state, if they succumb to the disease, or the recovered state, with different rates. Those who have recovered are assumed to be no longer susceptible to contracting the disease.

We introduce death rate *α* (*t*) as a time varying function, representing the proportion of infectious individuals who eventually die from the disease, by date. Those who eventually die transfer from the infected to the died state at a rate of *ρ*, representing the inverse of the time from becoming infectious until time of death. In our model, *ρ* is assumed to be constant over time. Those who eventually recover do so at the *γ*, representing the inverse of the time from becoming infectious until recovery. We will also later derive the effective reproduction number *Rep* (*t*), representing the average number of persons who are exposed to the disease by each infectious person, as a function of time.

Taking these factors into account, the system of equations of the proposed SEIRD model is given by::

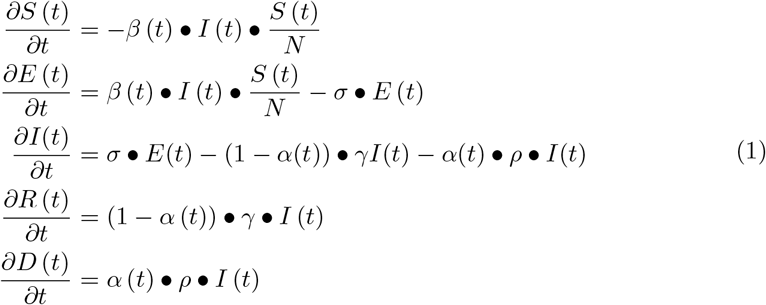

where:

*S* (*t*) = number of people in susceptible state at time t

*E* (*t*) = number of people in exposed, but uninfected at time t

*I* (*t*) = number of people in infectious state at time t

*D* (*t*) = number of people who have died at time t

*R* (*t*) = number of people who have recovered at time t N = total number of people

*β* (*t*) = transmission rate at time t

*s* = transformation rate from exposed to infectious, which is the reciprocal of the incubation period

*α* (*t*) = likelihood of eventual death of a person who is infected at time t

*γ* = transformation rate from infectious to recovered, which is the reciprocal of the recovery time

*ρ* = transformation rate from infectious to death

Changes in intervention policy, global events and medical care affect *α* (*t*) and *β* (*t*). While, in theory, these functions may change erratically as a consequence of discrete events, such as new public health measures, we hypothesize that such discrete events do not suddenly alter either function. Therefore, we seek to understand whether a simple continuous model, with a minimal set of parameters, might accurately represent historical data. For illustration, at the enactment of a new intervention policy, the public may not react immediately, and neither do the transmission parameters. The public will get used to the policy after a period of adaptation, and eventually the effective reproduction number will stabilize. In addition, the public responds to both government policies and communication about the disease. Communication comes from many, sometimes conflicting, sources. How the public at large absorbs and responds to such often confusing messages may be gradual.

A natural function to describe this pattern of change is the Sigmoid function.

Equation 2 is the general form of the Sigmoid function, where *k* determines the slope of the function and a determines the *x* value at the middle point (i.e., point of time when *y* = .5). A standard Sigmoid function with *k* = 1 and *a* = 0 is shown in Figure 1.

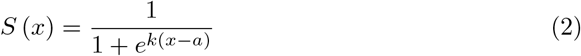

**Fig 1.**
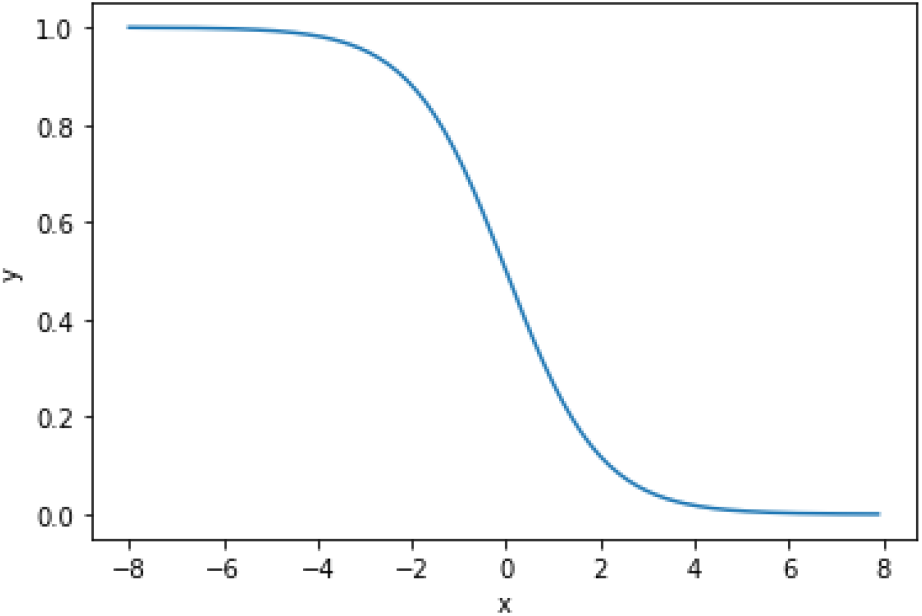
Standard Sigmoid function *k* = 1 and *a* = 0.

Thus, we define the function for transmission rate and death rate as:

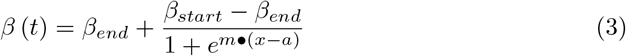

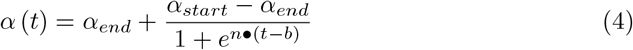

where:

*β*_*start*_ is the starting reproduction number

*β*_*end*_ is the ending reproduction number

*α*_*start*_ is the starting death rate, ranging from 0 to 1

*α*_*end*_ is the ending death rate, ranging from 0 to 1

*m, n, a, b* are the shape parameters

## Parameter Estimation and Model Fitting

Parameters in Eqs. 1 were estimated with the objective of minimizing the weighted summation of squared error between cumulative predicted and measured confirmed cases and the summation of squared error between cumulative predicted and cumulative confirmed deaths. Our analysis is based on the period from the day of first reported case in each state until 07/28/2020, across all 50 American states. For each state of the USA, we chose a start date of 4 days prior to the date of the first confirmed case. Four days was chosen based on a report by CDC [17], indicating the median incubation period is 4 days, ranging from 2 - 7 days.

Two methods were used for different sets of parameters, as described below. To estimate the shape parameters *m, n, a, b* and the starting/ending parameters *β*_*start*_, *β*_*end*_, *α*_*start*_, *α*_*end*_, we fit Eqs. 1 to the cumulative confirmed case numbers and the cumulative confirmed death numbers with the nonlinear least square method. Other parameters were derived from prior research.

## Parameters Derived from Prior Research

As mentioned in the CDC reports [17–19], the median incubation period is 4 days, ranging from 2 - 7. Among 305 hospitalized patients and 10,647 recorded deaths, the median time of hospitalization was 8.5 days and the median interval from illness onset to death was 10 days (IQR = 6 - 15 days). We assume the median hospitalization time is the median time for infectious people to stop being contagious. Hence, we set these parameters as the reciprocal of these time values: *σ* = 1*/*4, *γ* = 1*/*8.5, *ρ* = 1*/*10.

## Parameters Derived from Optimization

The remaining parameters are derived for each American state by optimizing the fit of the model to historical case and death data, where the objective is to minimize a weighted sum of daily squared error over the analysis period. We utilized a search algorithm that required initialization and a constrained search space, as explained below.

We define the model function *M* (*t*; [*β*_*start*_, *β*_*end*_, *m, a, α*_*start*_, *α*_*end*_, *n, b*]) : *t →R*^2^, where 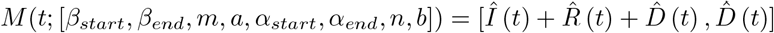 and the reported case number and death number at time t is [*Cases*(*t*), *Deaths*(*t*)].

Because it is unlikely for transmission and death rates to change drastically in a single day, we set upper bounds for m and n at 0.33 (meaning that rates do not suddenly change in less than three days) and initialize the search at 0.25. We permit the turning point of the sigmoid function to occur on any day in the timeline; we set *a, b* ∈ [0, 125], where 125 is the length of the period from March 1^*st*^ to July 28^*th*^, in days (as of March 1^*st*^ few states had reported cases). Prior research suggests that the initial effective reproduction number is around 3 [5], equivalent to a transmission rate of 0.75, which we use for initialization. Because transmission rates vary significantly among locations due to local conditions (such as crowding), we bound *β*_*start*_ ∈ [0.5, 7.5] and *β*_*end*_ ∈ [0, 2.5], thus permitting a wide range of results.

To summarize, the parameters set *P* = [*β*_*start*_, *β*_*end*_, *m, a, α*_*start*_, *α*_*end*_, *n, b*] is initialized as [0.75, 0.5, 0.25, 10, 0.4, 0.1, 0.25, 10]. Then the parameter optimization problem is:

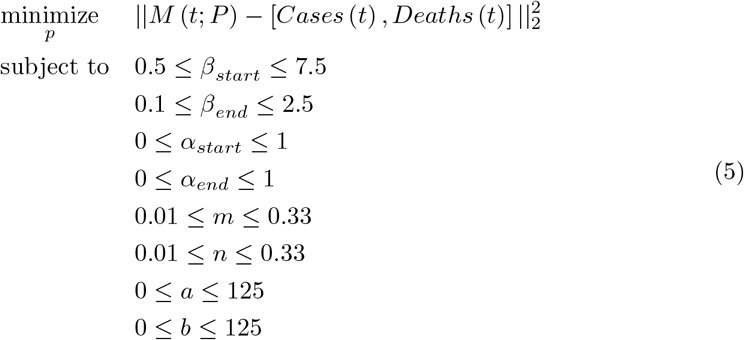

The number of reported deaths is smaller than the number of reported cases in all locations. Thus, treating errors in death estimation and case estimation the same will lead to underfitting of the death data, in preference to minimizing the errors in case data. Therefore, considering the accuracy of the reported death data and the fitting accuracy, we optimized a weighted sum of squared death and case data, multiplying w by deaths during the fitting process. The adjusted objective function is:

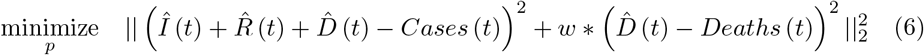

The parameters are estimated by solving the nonlinear constrained least-squares problem in Eqs. 6, utilizing the Levenberg–Marquardt algorithm (LMA). The LMA algorithm adaptively varies the parameter updates between the gradient descent update and the Gauss-Newton update and accelerates to a local minimum [20]. The LMA is implemented to our model fitting by the *lmfit* package in Python. In our analysis we utilized *w* = 20, to yield similar error percentages for deaths and cases.

## Data Limitations

We recognize that reported data are not the same as actual cases and actual deaths, which are unknowable. Daily confirmed cases are influenced by widely varying testing rates and policies, which change over time. At the beginning of the epidemic, the limited test kits were restricted to those who suffer from severe symptoms and those who are in a higher risk of exposure. Death data are likely to be more accurate, but they too can suffer from reporting errors, due to how deaths are attributed to COVID-19 (or not), the timing of filing reports and the general accuracy of reporting. For these reasons, our model represents estimation of reported data rather than the actual (but unknown) number of cases and number of deaths.

Reporting has also shown a consistent day-of-week variation across many locations, with weekend data differing from weekday data. This variation is more likely the consequence of different patterns of healthcare staffing, and differences in how patients present for testing by day of week, rather than differences in disease transmission by day of week. To smooth out these effects, we model the moving 7-day average data instead of the daily reported data

## Results

### Model Accuracy

The first case of COVID-19 in the United States was reported on January 20, 2020 [21]. As of July 28, 2020, a total of 4,459,2869 cases and 151,711 deaths had been reported across the states and territories of America [22]. We fit the model with the dataset of 7-day moving average cases and deaths for the 50 states, provided by the COVID-19 tracking project lead by The Atlantic (derived from the Center for Disease Control), for the period from the date of the first reported cases to July 28^*th*^. The fitting accuracy across all states is presented in Figure 2, measured by the relative root mean square error (RRMSE):

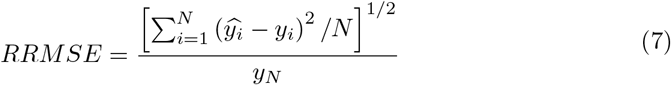

where *y*_*N*_ is the case/death number on the *N*_*t*__*h*_ day. The fitting accuracy of the reported case number ranges from 0.54% to 7.34% and of the reported death number ranges from 0.29% to 7.28%. The average and median RRMSEs for deaths are 1.61% and 1.33%; for cases, the average and median values are 2.30% and 1.88%. RRMSE fell below 5% by both measures for all states except Hawaii, Louisiana, Montana and Wyoming. Figure 2 shows that the proposed SEIRD model with time-dependent transmission rate and death rate captured the pattern of the transmission dynamics well across most states with only 8 fitted parameters.

**Fig 2.**
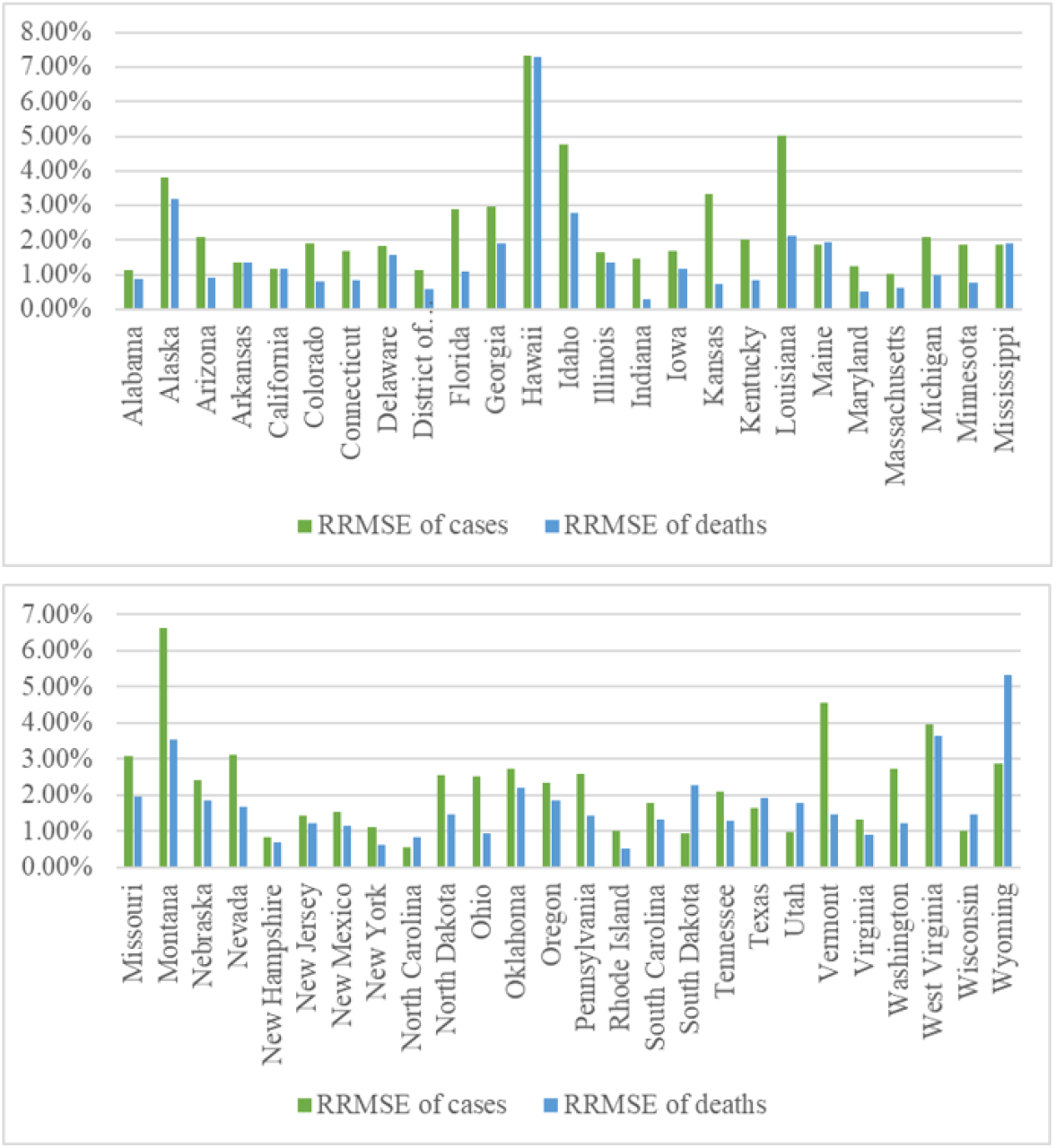
Fitting accuracy of the cases and fatality across all states.

Figure 3 and 4 show the specific fitting results for cases and deaths by day for the two states with the largest number of cases (New York and California) as well as two other states for which the fit is less accurate (Florida and Hawaii). For New York and California, the fitting results almost coincide with the CDC data. Examining Florida and Hawaii, the CDC data follows a pattern of two phases, which is not as well captured by our model. Especially for Hawaii, the curve flattened for a period and then rose. As discussed later, our model characterizes the transmission dynamic for a period with one phase, i.e. the curve should become flat at most once.

**Fig 3.**
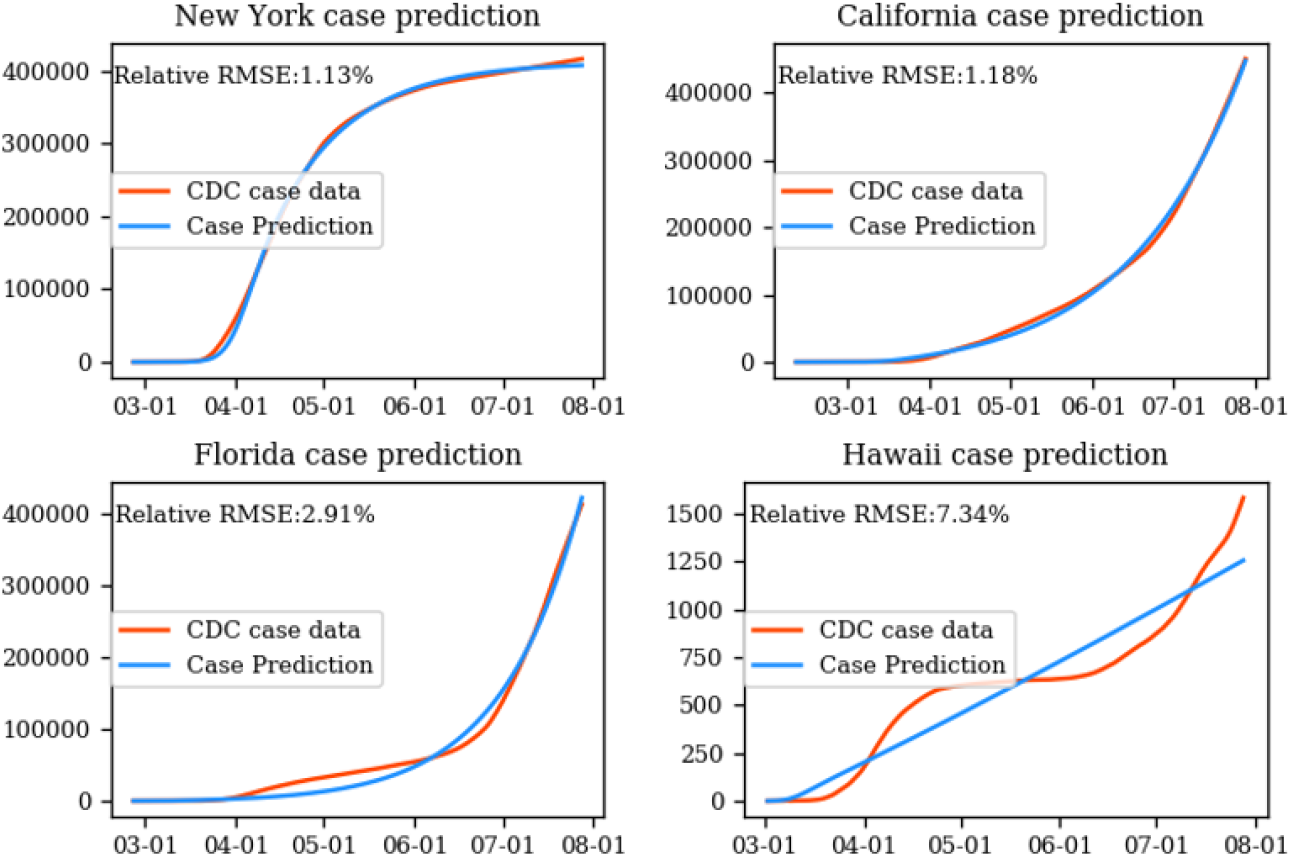
Fitting results of case number for New York, California, Florida and Hawaii.

**Fig 4.**
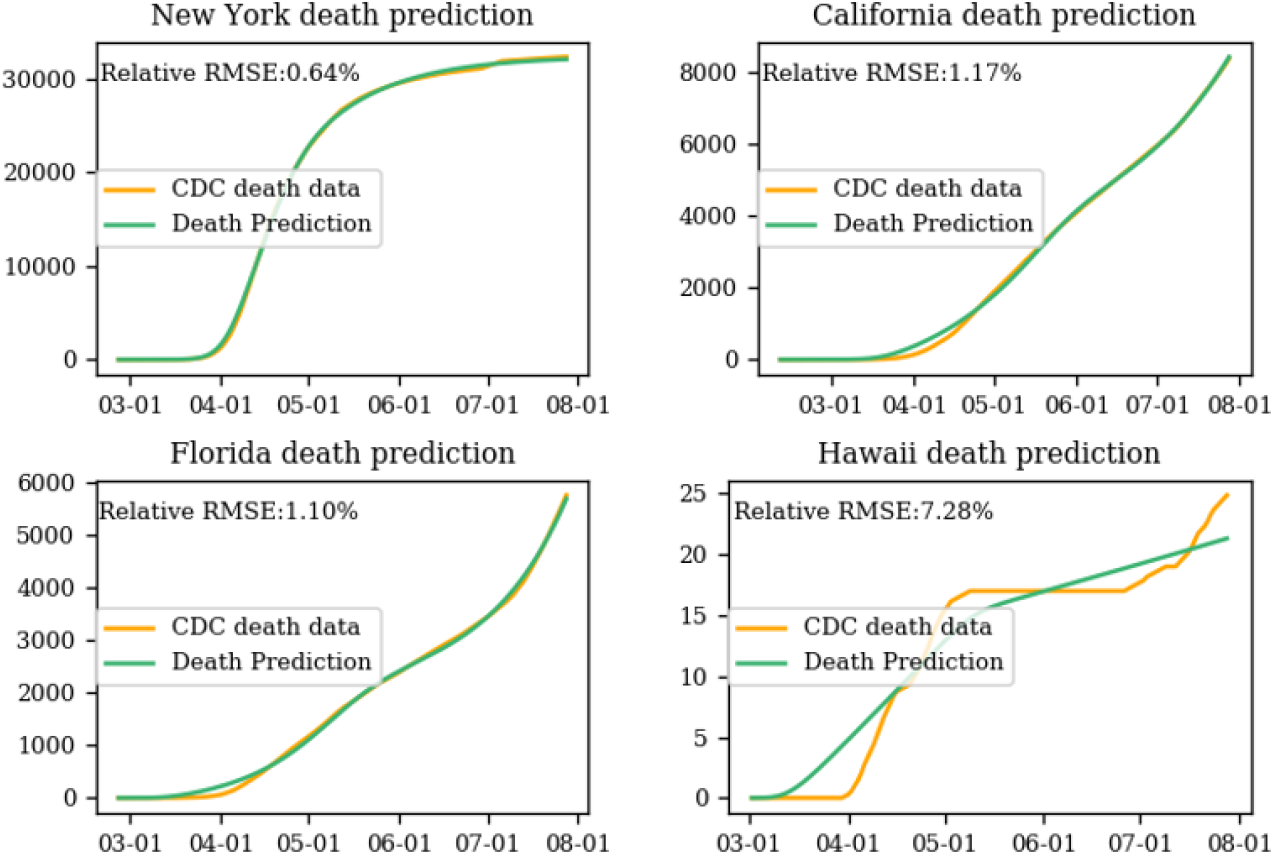
Fitting results of death number for New York, California, Florida and Hawaii.

## Effective Reproduction Number Calculation and Trends

Effective reproduction number at any time t, which we define as *Rep* (*t*), is the average number of people in a population who are infected per infectious case, where everyone is susceptible to the disease. *Rep* (*t*) measures the transmission potential of infectious diseases [23]. When *Rep* (*t*) *>* 1, the rate of new cases will increase over time, until the population loses susceptibility to the disease. When *Rep* (*t*) *<* 1, the rate of new cases will decline over time.

*Rep* (*t*) can be estimated with the next generation matrix method [24]. We define X as the vector of infected class (i.e. E, I) and Y as the vector of uninfected class (i.e. S, R, D). Let 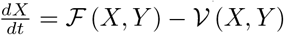, where ℱ(*X, Y*) is the vector of new infection rates (flows from Y to X) and (*X, Y*) is the vector of all other rates. Then for our model, the next generation matrix is expressed as

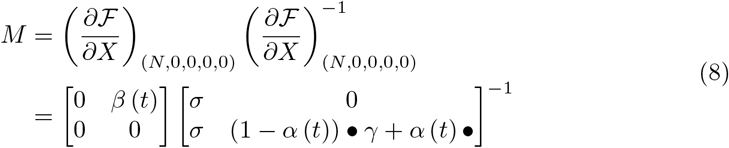

Then the effective reproduction number is the spectral radius of M, expressed as:

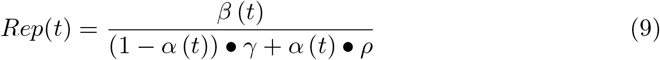

At the beginning of the epidemic, *Rep* (*t*) reflects the natural transmissibility of COVID-19, i.e. the basic reproduction number *R*0 in the absence of intervention. With the evolution of the epidemic, *Rep* (*t*) changes dynamically, as do the transmission rate *β* (*t*) and death rate *α* (*t*), which are influenced by both the intervention policy and population immunity. Figures 5 and 6 show the fitted *Rep* (*t*) at the start of the epidemic across all states and fitted *Rep* (*t*) on July 28^*th*^. We see that *Rep* (*t*) ranges from 1.27 to 16.49, with a median value of 2.87. It should be kept in mind that this optimal fit is a reflection of the reported data on cases. Increasingly aggressive testing may make it appear that *Rep* (*t*) grows faster than the actual (unknown) number of cases.

**Fig 5.**
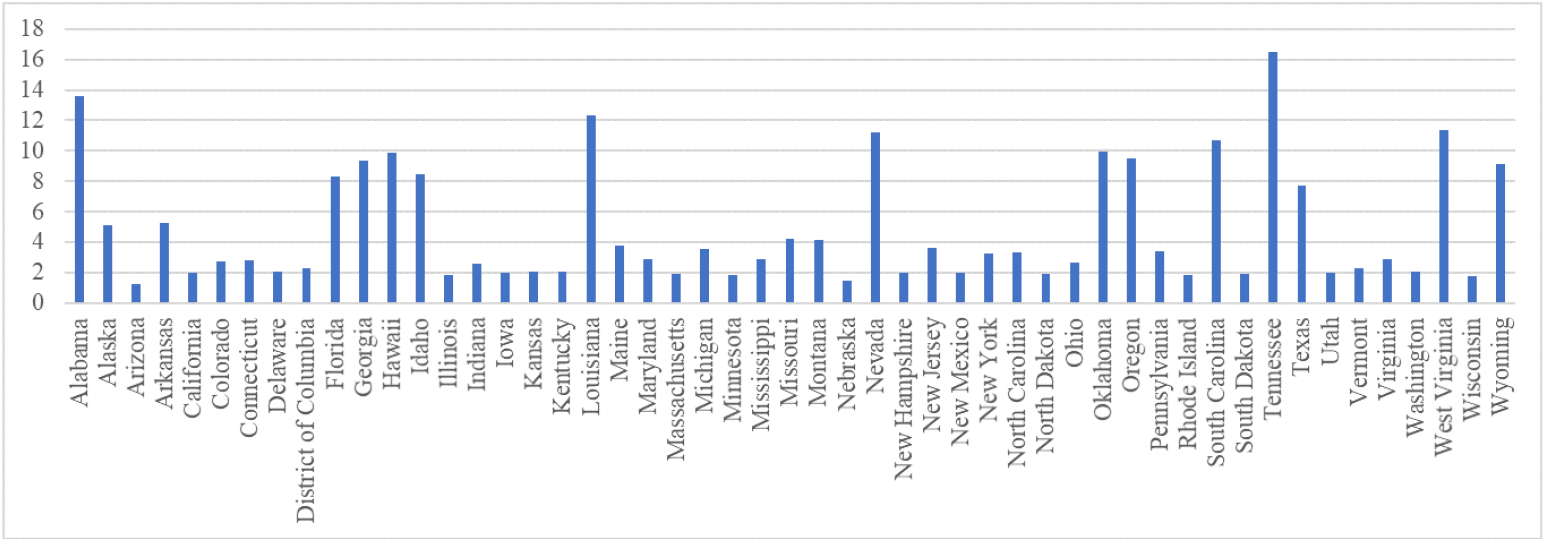
Fitted *Rep* (*t*) at the start of the epidemic This figure shows the reproduction number on the date of first reported case across all states

**Fig 6.**
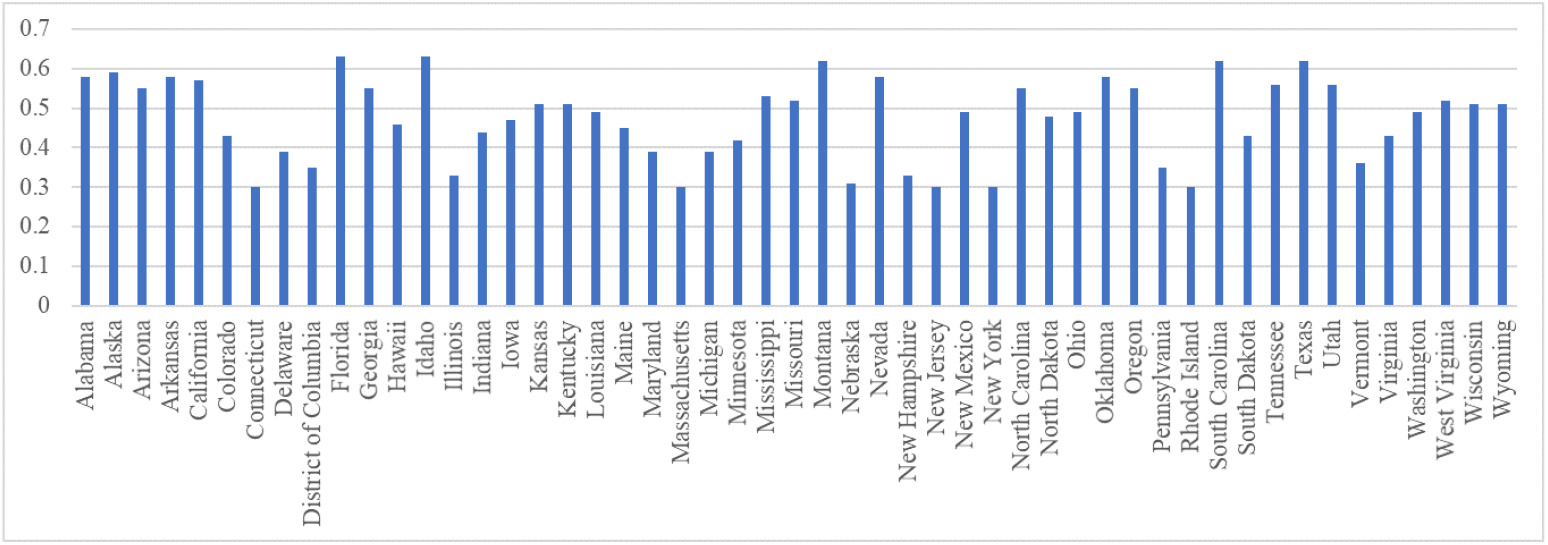
Fitted *Rep* (*t*) on July 28This figure shows the reproduction number on July 28 across all states

For illustration, Figure 7 shows our estimated history of *Rep* (*t*) for New York, California, Florida and Hawaii. Time 0 in these graphs is the day of the first reported case, which varies from state to state. In these cases, the effective reproduction number both stabilized and became smaller than 1 with time, with the change occurring over a period of 10 to 30 days.

**Fig 7.**
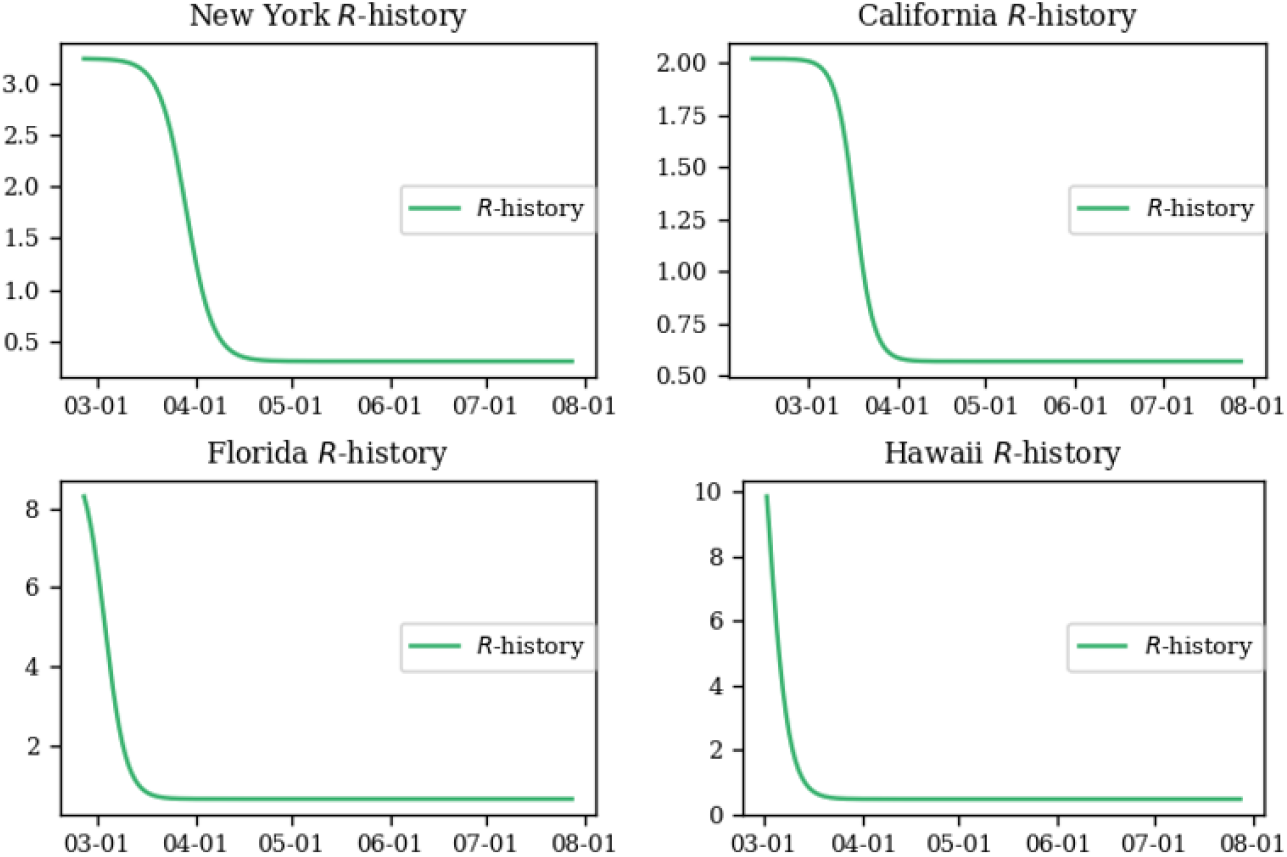
History of effective reproduction number for New York, California, Florida and Hawaii.

As noted, in the early stages of an epidemic, the reproduction number may seem particularly large not only because the disease spreads rapidly but also because the rate of testing is increasing. In this sense, the estimated reproduction number is a reflection of both changes in the data collection process and the actual spread of disease.

## Death Rate Trends

Death rate is another measure that shows the change in virus outcomes over time, reflecting the health system’s ability to deal with the flood of infected people. Figure 8 provides examples. From the historical plot, we see the hardest-hit states, like New York and Florida, experienced a much higher death rate in the early stage than the average 3% death rate in the United States. The relatively high death rate could be caused by the lack of effective medical treatment and hospital overload. It could also reflect limited testing of patients, whereby only the sickest patients were recorded as cases. With improvement of medical treatment, and perhaps increased testing, the death rate per confirmed case for most states decreased to a stable value.

**Fig 8.**
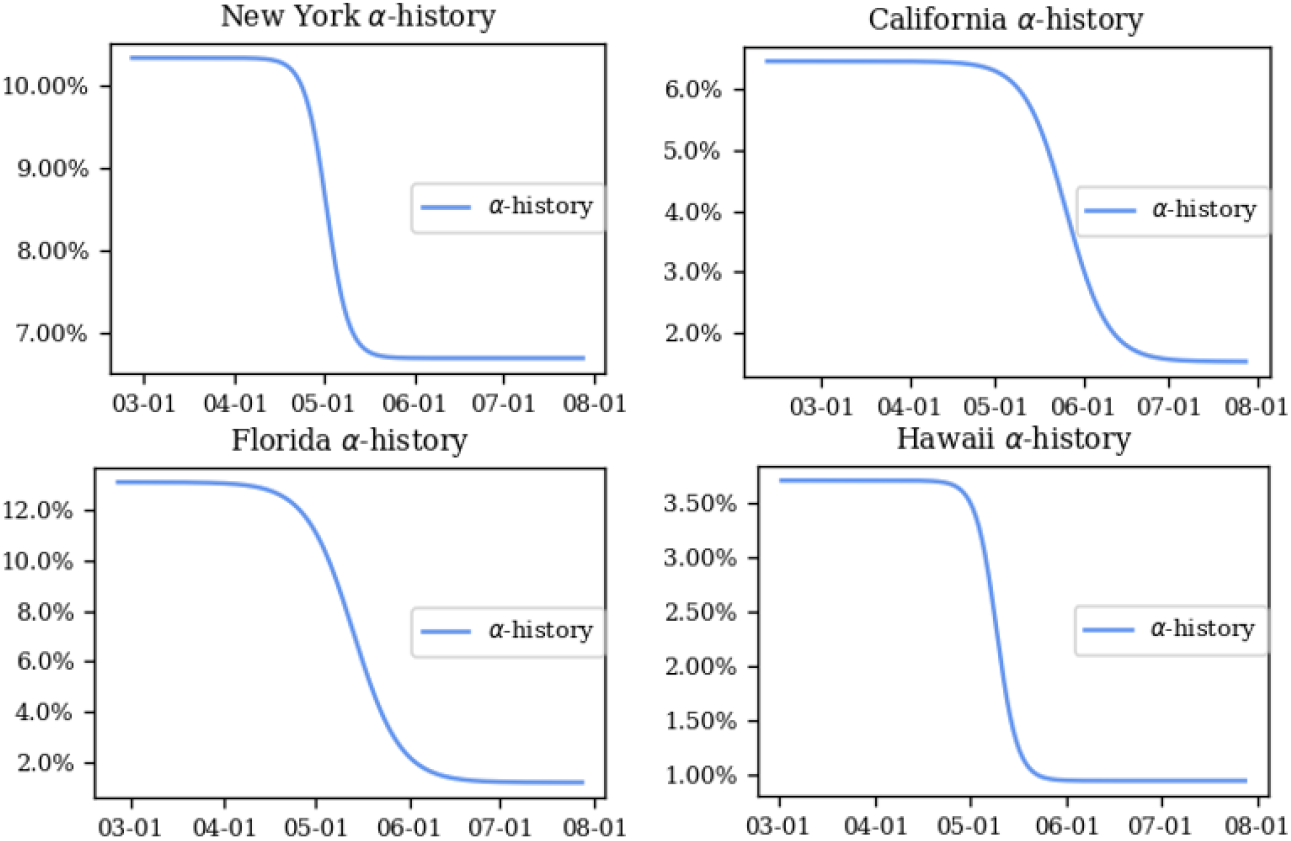
History of death rate for New York, California, Florida and Hawaii.

## Multi-Phase Model

Our model as presented fits reported data for cases and deaths within 2% error in most states. However, because it is premised on the assumption that transmission rates do not at first go down, and then later go up, it needs to be modified for states that exhibit multiple waves of the disease. Data for Hawaii – for which the model has the poorest fit – are most indicative of this pattern.

For such locations, we propose an alternate multi-phase model. The Hawaii Department of Health announced the first positive case on Oahu, Hawaii, on March 6^*th*^ and then enacted a stay at home order on March 25^*th*^. From April 19^*th*^ to May 7^*th*^, the case curve flattened. The state announced on May 7^*th*^ that Hawaii would embark on the first phase of reopening. The data reflect a second wave of coronavirus commencing on or about May 7^*th*^.

We divide the Hawaii timeline into two periods, the first from March 6^*th*^ until May 7^*th*^, and the second from May 7^*th*^ to July 28^*th*^. We fit the first stage with the initialization of only one exposed people at the start. To initialize the second phase, we use the predicted number of exposed people, infectious people and recovered people from the first phase, combined with the reported deaths as of May 7^*th*^. With this modification, the RRMSE for cases declines below 2.5% and the RRMSE for deaths declines below 2.7%. The fitting results in Figure 9 show that our two-phase model captures the transmission pattern more precisely than the single-phase model.

**Fig 9.**
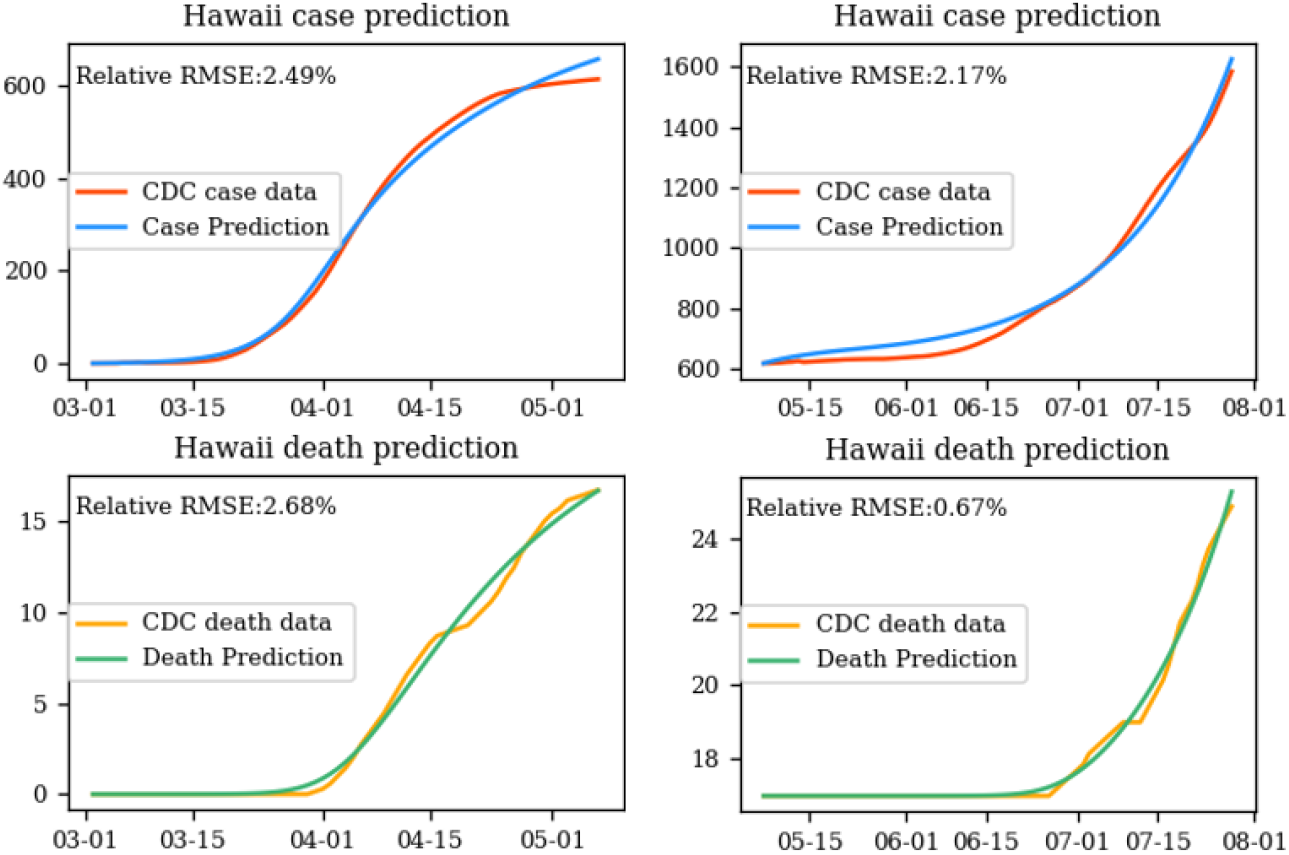
Fitting results for the two phases Hawaii. The first phase (left figures) starts from March 6 to May 7; The second phase (right figures) starts from May 7 to July 28.

The history of effective reproduction number and death rate are shown in Figure 10.

**Fig 10.**
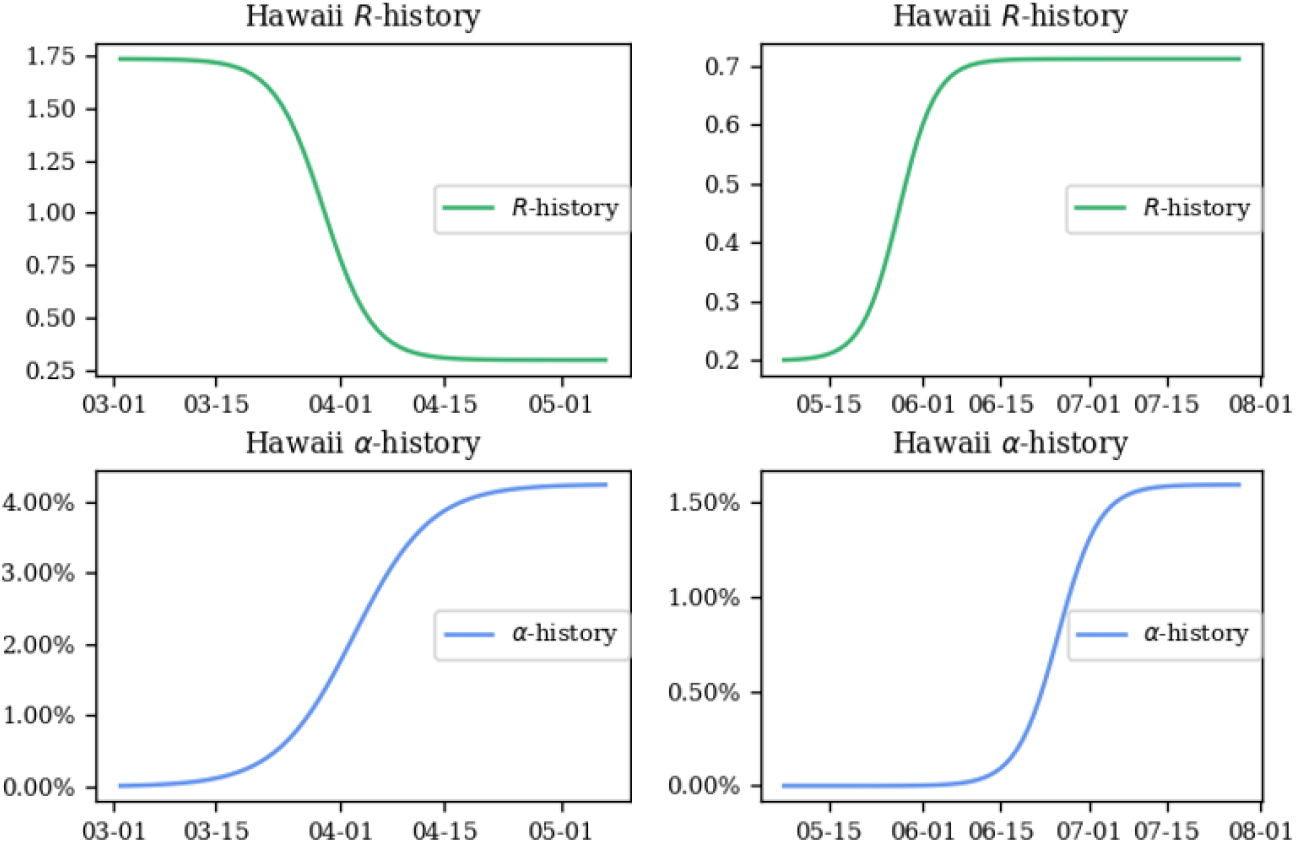
Historical results of the effective reproduction number and death rate. The first phase (left figures) starts from March 6 to May 7; The second phase (right figures) starts from May 7 to July 28.

The first phase showed a decline in the reproduction number after the initial announcement of the stay-at-home order. However, with the reopening, the reproduction number increased, explaining increases in case rates. Death rates, by contrast, exhibit a peculiar behavior, increasing over time in each phase, with a discontinuity when transitioning from the first phase to the second. Beyond exhibiting two phases, Hawaii has a small number of deaths, with no deaths occurring in the transition period between phases. We surmise that the function, while representing the data well, is peculiar because of the unusual pattern in deaths within Hawaii.

## Conclusions

We have developed an extension of the SEIRD model that represents changing transmission and death rates over time as a continuous Sigmoid function, under the hypothesis that these rates change gradually, rather than immediately upon implementation of public health policies or treatments. The model fits historical data for the United States well for most states. Those with poorer fits exhibit patterns of multiple waves of the disease. Using Hawaii as an example, a multi-phase extension of the model provides a more accurate fit, where the transition from one phase to the next is defined by a change in public health policy.

An advantage of the model is that it is defined by a small set of parameters. Thus, it provides an efficient method for quantifying differences among regions in the spread and outcomes of the disease. In the future, we intend to examine the predictive value for the model, taking into consideration ranges of uncertainty in model estimates. By examining historical trends, we can understand how variations in simple parameters can lead to fewer or more cases and deaths. In the future, we will develop multi-region extensions of the model, which permit representation of spread of disease from one region to another, or perhaps within sub-regional groups.

Our research is premised on the method of modeling case and death data as they are reported. We recognize that the true number of cases may differ from reported values, as might the number of deaths, both of which are unknown. The variations from state to state reflect in part the actual spread and outcomes of disease, as well as the extent to which cases are detected and reported, as well as how deaths have been classified.

## Data Availability

All data are fully available without restriction

## Supporting information

**S1 File. Fittingresults US**.**txt** Results of the estimated parameter values across 50 states in the US.

**S2 File. Statesreport**.**csv** Fitting accuracy of the case and death number across 50 states in the US.

## Author Contributions

**Conceptualization:** Mingdong Lyu, Randolph Hall

**Formal analysis:** Mingdong Lyu, Randolph Hall

**Funding acquisition:** Randolph Hall, Mingdong Lyu

**Investigation:** Mingdong Lyu, Randolph Hall

**Project administration:** Randolph Hall

**Methodology:** Mingdong Lyu

**Writing - original draft:** Randolph Hall, Mingdong Lyu

**Writing - review & editing:** Randolph Hall, Mingdong Lyu

## References

1. Fang Y, Nie Y, Penny M. Transmission dynamics of the COVID-19 outbreak and effectiveness of government interventions: A data-driven analysis. Journal of medical virology. 2020;92(6):645–659.

2. Tesini BL. Coronaviruses and Acute Respiratory Syndromes (COVID-19, MERS, and SARS) - Infectious Diseases. Merck Manuals; 2020. Available from: https://www.merckmanuals.com/professional/infectious-diseases/respiratory-viruses.

3. Backer JA, Klinkenberg D, Wallinga J. Incubation period of 2019 novel coronavirus (2019-nCoV) infections among travellers from Wuhan, China, 20–28 January 2020. Eurosurveillance. 2020;25(5):2000062.

4. Zhao S, Lin Q, Ran J, Musa SS, Yang G, Wang W, et al. Preliminary estimation of the basic reproduction number of novel coronavirus (2019-nCoV) in China, from 2019 to 2020: A data-driven analysis in the early phase of the outbreak. International journal of infectious diseases. 2020;92:214–217.

5. Liu Y, Gayle AA, Wilder-Smith A, Rocklöv J. The reproductive number of COVID-19 is higher compared to SARS coronavirus. Journal of Travel Medicine. 2020 02;27(2). Taaa021. Available from: https://doi.org/10.1093/jtm/taaa021.

6. New York State Task Force on Life & the Law NYSDoH. VENTILATOR ALLOCATION GUIDELINES; 2015. November.

7. Rosenthal BM, Goldstein J. N.Y. May Need 18,000 Ventilators Very Soon. It Is Far Short of That. The New York Times; 2020. Available from: https://www.nytimes.com/2020/03/17/nyregion/ny-coronavirus-ventilators.html.

8. COVID I, Murray CJ, et al. Forecasting COVID-19 impact on hospital bed-days, ICU-days, ventilator-days and deaths by US state in the next 4 months. medRxiv. 2020;.

9. Moghadas SM, Shoukat A, Fitzpatrick MC, Wells CR, Sah P, Pandey A, et al. Projecting hospital utilization during the COVID-19 outbreaks in the United States. Proceedings of the National Academy of Sciences. 2020;117(16):9122–9126.

10. Zhao S, Musa SS, Lin Q, Ran J, Yang G, Wang W, et al. Estimating the unreported number of novel coronavirus (2019-nCoV) cases in China in the first half of January 2020: a data-driven modelling analysis of the early outbreak. Journal of clinical medicine. 2020;9(2):388.

11. Nishiura H, Kobayashi T, Miyama T, Suzuki A, Jung Sm, Hayashi K, et al. Estimation of the asymptomatic ratio of novel coronavirus infections (COVID-19). International journal of infectious diseases. 2020;94:154–155.

12. Radulescu A, Cavanagh K. Management strategies in a SEIR model of COVID 19 community spread. arXiv preprint arXiv:200311150. 2020;.

13. Luo G, McHenry ML, Letterio JJ. Estimating the prevalence and risk of COVID-19 among international travelers and evacuees of Wuhan through modeling and case reports. PloS one. 2020;15(6):e0234955.

14. Zhao S, Zhuang Z, Ran J, Lin J, Yang G, Yang L, et al. The association between domestic train transportation and novel coronavirus (2019-nCoV) outbreak in China from 2019 to 2020: a data-driven correlational report. Travel medicine and infectious disease. 2020;33:101568.

15. Godio A, Pace F, Vergnano A. SEIR Modeling of the Italian Epidemic of SARS-CoV-2 Using Computational Swarm Intelligence. International Journal of Environmental Research and Public Health. 2020;17(10):3535.

16. Loli Piccolomini E, Zama F. Monitoring Italian COVID-19 spread by a forced SEIRD model. PloS one. 2020;15(8):e0237417.

17. Guan Wj, Ni Zy, Hu Y, Liang Wh, Ou Cq, He Jx, et al. Clinical characteristics of coronavirus disease 2019 in China. New England journal of medicine. 2020;382(18):1708–1720.

18. Gold JA. Characteristics and clinical outcomes of adult patients hospitalized with COVID-19—Georgia, March 2020. MMWR Morbidity and mortality weekly report. 2020;69:545–550.

19. Wortham JM. Characteristics of persons who died with COVID-19—United States, February 12–May 18, 2020. MMWR Morbidity and mortality weekly report. 2020;69:923–929.

20. Moré JJ. The Levenberg-Marquardt algorithm: implementation and theory. In: Numerical analysis. Springer; 1978. p. 105–116.

21. Holshue ML, DeBolt C, Lindquist S, Lofy KH, Wiesman J, Bruce H, et al. First Case of 2019 Novel Coronavirus in the United States. New England Journal of Medicine. 2020;382(10):929–936.

22. The COVID Tracking Project. The Atlantic; 2020. Available from: https://covidtracking.com.

23. Nishiura H, Chowell G. The effective reproduction number as a prelude to statistical estimation of time-dependent epidemic trends. In: Mathematical and statistical estimation approaches in epidemiology. Springer; 2009. p. 103–121.

24. Van den Driessche P. Reproduction numbers of infectious disease models. Infectious Disease Modelling. 2017;2(3):288–303.

